# Community Pharmacists’ Challenges with Non-Codeine OTC Cough Syrup Abuse Among Nigerian Youths

**DOI:** 10.1101/2025.10.22.25338550

**Authors:** Samuel David Chinonyerem

## Abstract

Non-prescription cough syrups containing active ingredients like diphenhydramine and dextromethorphan are widely available in Nigeria without a prescription. However, their misuse among youths aged 15–35 is a growing public health issue. This study surveyed 45 community pharmacists in Lagos, Ogun, Ondo, Rivers, and Abuja to understand their challenges in addressing this problem (response rate: 75%, 45 out of 60 invited). Results show that 66.7% of pharmacists often or sometimes see youths buying large quantities of these syrups, with diphenhydramine linked to 77.8% of suspected abuse cases. Common signs of misuse include purchasing multiple bottles (66.7%) and suspicious behaviour during sales (66.7%). Pharmacists face barriers such as regulations allowing over-the-counter (OTC) sales (44.4%), fear of losing customers (11.1%), and feeling they lack authority to refuse sales (22.2%). While 44.4% regularly counsel youths on misuse risks, none reported adverse effects to the National Agency for Food and Drug Administration and Control (NAFDAC). Reasons for not reporting include unclear processes (33.3%) and lack of time (22.2%). Despite challenges, 55.6% of pharmacists feel confident identifying abuse signs. Suggested solutions include stricter regulations (77.8%) and mandatory counselling for young buyers (66.7%). This study highlights the need for better regulations, improved drug monitoring, and more support for pharmacists to tackle this public health challenge.

## Introduction

The misuse of over-the-counter (OTC) medications, particularly non-codeine cough syrups, is a growing concern among Nigerian youths (Gobir et al., 2021). After the 2018 NAFDAC ban on codeine-containing syrups due to widespread abuse, non-codeine alternatives like diphenhydramine (a sedating antihistamine) and dextromethorphan (a cough suppressant with dissociative effects at high doses) have become common targets for misuse (Agada et al., 2021). These substances, when abused, can cause serious health issues such as dizziness, hallucinations, seizures, addiction, and mental health problems, especially when mixed with alcohol or soft drinks (Akunne et al., 2025).

Community pharmacists, often the first point of contact for these purchases, are in a key position to address this issue (Abubakar et al., 2022). However, they face challenges like navigating loose regulations, balancing business interests, and lacking tools to monitor misuse effectively. This study examines these challenges through a survey of pharmacists in selected Nigerian states. It explores signs of abuse, operational difficulties, reporting practices, and potential solutions to help develop better strategies to prevent youth drug misuse in Nigeria (United Nations Office on Drugs and Crime, 2021).

## Methods

### Study Design and Participants

An online survey was conducted in October 2025 to gather insights from community pharmacists in Nigeria. The survey included 45 actively practicing pharmacists, primarily from Lagos (82.2%), with others from Ogun (8.9%), Ondo (2.2%), Rivers (2.2%), and Abuja (4.4%). Only active pharmacists were included to ensure reliable and relevant responses.

### Data Collection

A structured questionnaire with 15 questions was created to explore pharmacists’ experiences. It covered their background, observations of misuse, challenges, interactions with youths, identification and reporting of adverse effects, confidence levels, and suggestions for improvement. The questionnaire included multiple-choice and open-ended questions to capture a wide range of insights. Hosted on Google Forms, the survey ensured confidentiality and was completed voluntarily to encourage honest responses.

### Data Analysis

Responses were analyzed to summarize key findings. Counts and percentages were used for multiple-choice questions. Relationships, such as years of experience and frequency of counselling, were examined to identify trends. Results were reviewed to understand their implications for drug monitoring and highlight areas for further research. Ethical standards were followed, with participant consent obtained and data aggregated to protect privacy.

## Results

### Demographic Profile

The pharmacists had an average of 5.6 years of experience (range: 2–10 years). Daily customer volumes varied as shown in Table 1.

**Table 1:**
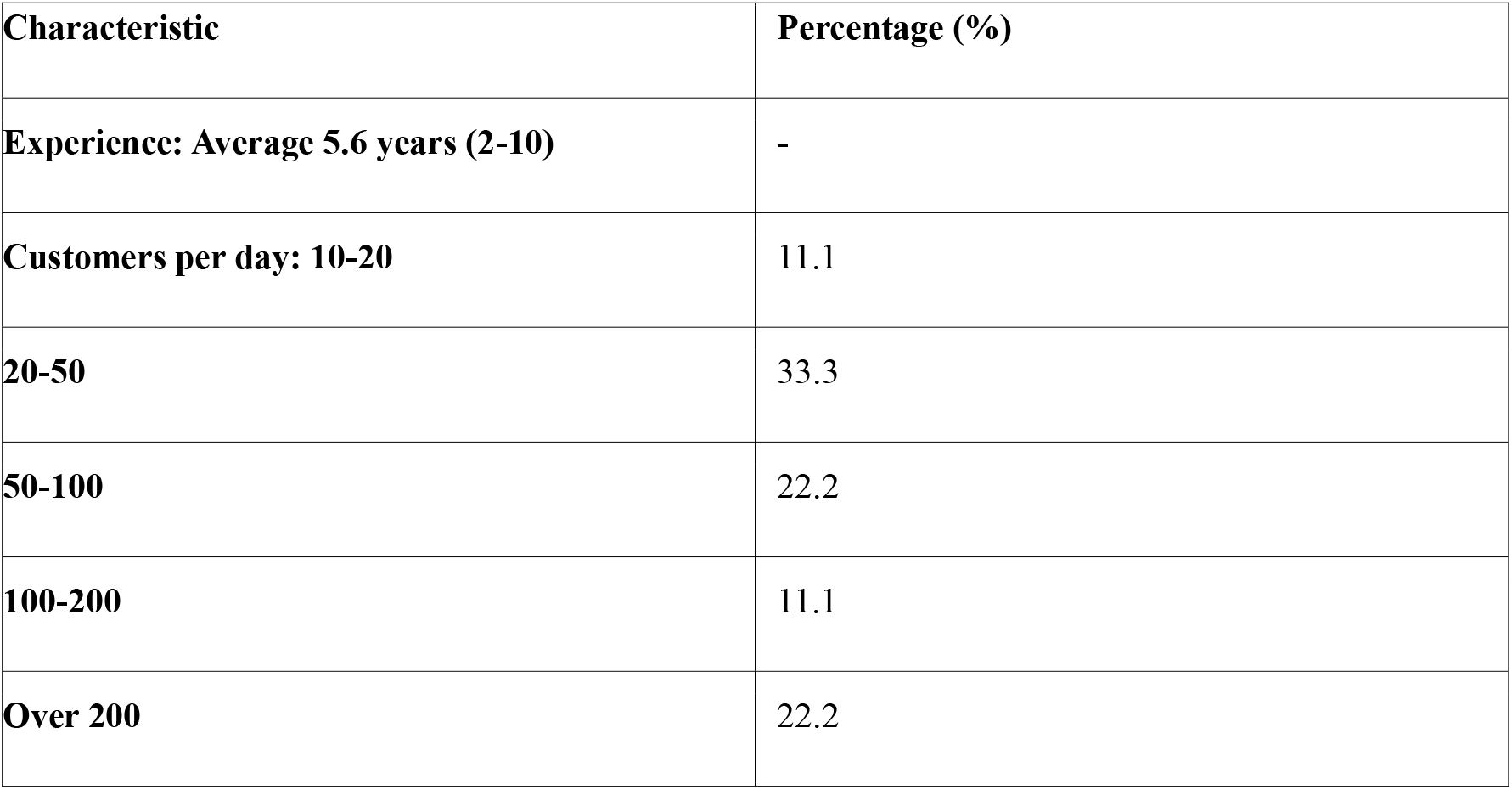
Demographic Profile of Pharmacists.

### Observations of Abuse

Most pharmacists (44.4%) reported frequent (weekly) purchases of non-codeine OTC cough syrups by youths, while 22.2% observed this monthly, and 33.3% saw it rarely or never. Diphenhydramine was the most commonly misused substance (77.8%), followed by dextromethorphan (11.1%) and mixed formulations (11.1%). Signs of misuse included frequent purchases (77.8%), buying multiple bottles (66.7%), suspicious behaviour (66.7%), and mixing syrups with alcohol or soda (66.7%). See Table 2 for details.

**Table 2:** Observations of Abuse.

| **Observation** | **Percentage (%)** |
| --- | --- |
| **Frequency: Weekly** | 44.4 |
| **Monthly** | 22.2 |
| **Rarely/Never** | 33.3 |
| **Misused Substance:**<br>**Diphenhydramine** | 77.8 |
| **Dextromethorphan** | 11.1 |
| **Mixed** | 11.1 |
| **Signs: Frequent purchases** | 77.8 |
| **Multiple bottles** | 66.7 |
| **Suspicious behaviour** | 66.7 |
| **Mixing with alcohol/soda** | 66.7 |

### Challenges in Prevention

Pharmacists identified several barriers: OTC regulations allowing sales without prescriptions (44.4%), lack of authority to refuse sales (22.2%), fear of losing customers (11.1%), fear of harm (11.1%), or a combination of these issues (11.1%). See Table 3.

**Table 3:**
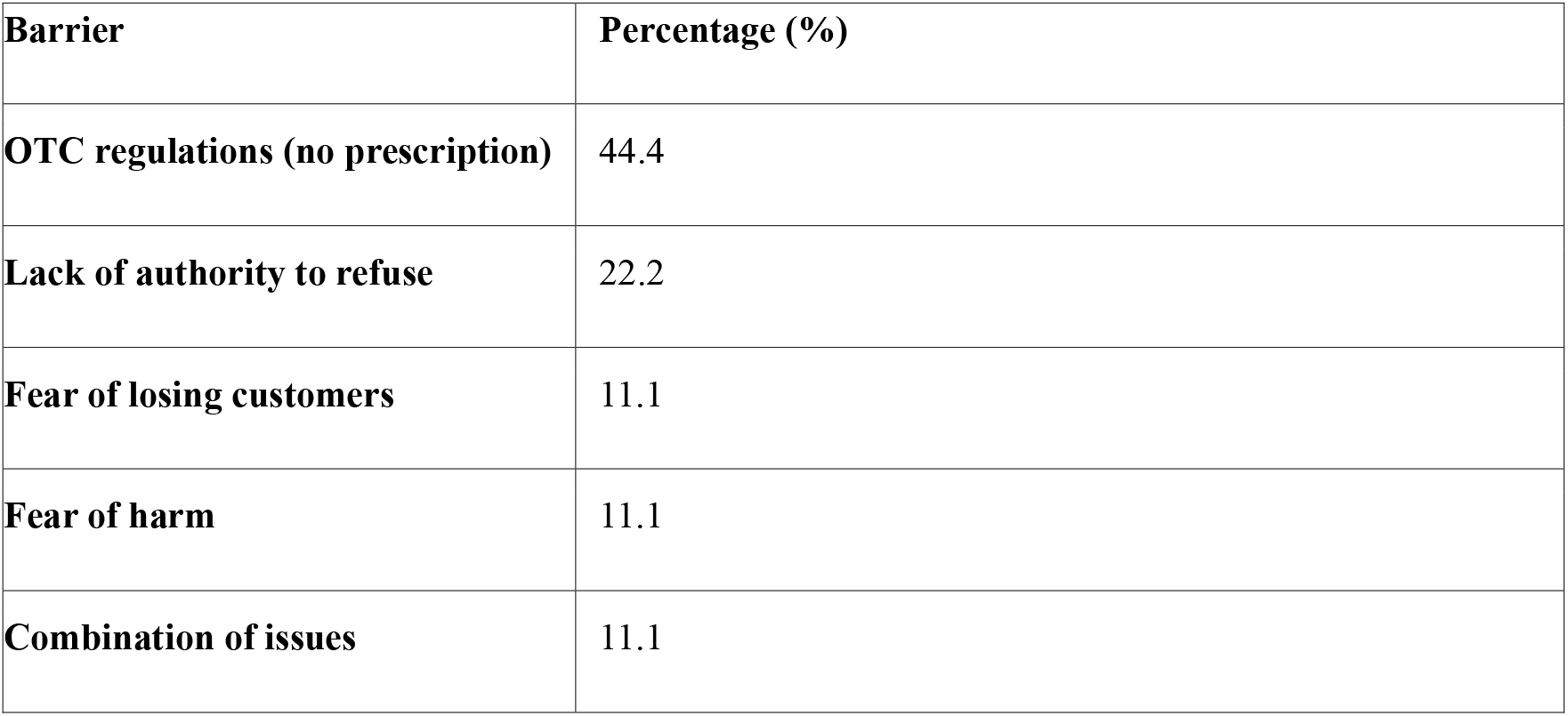
Challenges in Prevention.

| **Barrier** | **Percentage (%)** |
| --- | --- |
| **OTC regulations (no prescription)** | 44.4 |
| **Lack of authority to refuse** | 22.2 |
| **Fear of losing customers** | 11.1 |
| **Fear of harm** | 11.1 |
| **Combination of issues** | 11.1 |

### Counselling and Intervention Practices

Most pharmacists (44.4%) always counsel youths on misuse risks, 44.4% do so sometimes, and 11.1% rarely counsel. Barriers to counselling include lack of time (33.3%), perceived lack of authority (11.1%), uneducated customers (11.1%), youths obtaining syrups elsewhere (11.1%), fear of attack (11.1%), and lack of proof of misuse (11.1%). See Table 4.

**Table 4:** Counselling Practices and Barriers.

| **Practice/Barrier** | **Percentage (%)** |
| --- | --- |
| **Always counsel** | 44.4 |
| **Sometimes counsel** | 44.4 |
| **Rarely counsel** | 11.1 |
| **Barriers: Lack of time** | 33.3 |
| **Lack of authority** | 11.1 |
| **Uneducated customers** | 11.1 |
| **Obtain elsewhere** | 11.1 |

| Practice/Barrier | Percentage (%) |
| --- | --- |
| **Fear of attack** | 11.1 |
| **Lack of proof** | 11.1 |

### Adverse Drug Reaction (ADR) Suspicion and Reporting

While 33.3% suspected adverse effects from misuse, none reported these to NAFDAC. Reasons included unclear reporting processes (33.3%), lack of time (22.2%), fear of legal issues (11.1%), belief that reporting would not help (11.1%), or other reasons (22.2%). Most pharmacists (55.6%) felt very confident in spotting abuse, while 44.4% felt moderately confident. See Table 5.

**Table 5:** ADR Reporting and Confidence.

| Aspect | Percentage (%) |
| --- | --- |
| **Suspected ADRs** | 33.3 |
| **Reported to NAFDAC** | 0.0 |
| **Reasons: Unclear processes** | 33.3 |
| **Lack of time** | 22.2 |
| **Fear of legal issues** | 11.1 |
| **No help** | 11.1 |
| **Other** | 22.2 |
| **Confidence: Very confident** | 55.6 |
| **Moderately confident** | 44.4 |

### Proposed Strategies

Pharmacists suggested stricter OTC regulations (77.8%), mandatory counselling for young buyers (66.7%), training to identify abuse (55.6%), and user-friendly reporting apps (44.4%). Other ideas included community outreach, requiring prescriptions for high-risk syrups, public awareness campaigns, and closer collaboration with NAFDAC. See Table 6.

**Table 6:** Proposed Strategies.

| Strategy | Percentage (%) |
| --- | --- |
| Stricter OTC regulations | 77.8 |
| Mandatory counselling | 66.7 |
| Training to identify abuse | 55.6 |
| User-friendly apps | 44.4 |

## Discussion

This study confirms that non-codeine OTC cough syrup abuse is widespread among Nigerian youths, with diphenhydramine being the most commonly misused substance, similar to trends observed globally (Sessa et al., 2021; Garba et al., 2023). The high frequency of abuse (66.7% of pharmacists see it weekly or monthly) may have increased since the 2018 codeine ban, potentially leading to unreported health issues (United Nations Office on Drugs and Crime, 2021).

Current OTC regulations enable legitimate use but also facilitate abuse (National Agency for Food and Drug Administration and Control, 2025). Pharmacists feel limited by business pressures and lack of authority to refuse sales, highlighting weaknesses in the system (Abubakar et al., 2022). The absence of adverse effect reports to NAFDAC, despite 33.3% suspecting issues, is concerning and stems from unclear processes and time constraints (Aina et al., 2023). This gap hides the true scale of the problem and delays regulatory action (Abdulaziz et al., 2025).

Pharmacists with more experience (over 5 years) were more likely to counsel youths (60% vs. 30% for those with less experience), suggesting training could improve interventions. However, inconsistent counselling (44.4% only counsel sometimes) indicates a need for better support and resources.

These findings point to serious public health risks, including increased healthcare costs and undetected harm to youths (Akunne et al., 2025). Solutions like prescription-only status for high-risk syrups, user-friendly reporting tools, and collaboration with NAFDAC could help address the issue (National Agency for Food and Drug Administration and Control, 2025). Study limitations include reliance on self-reported data, potential bias, and coverage of only select states.

## Conclusion

Community pharmacists in Nigeria face significant challenges in addressing non-codeine OTC cough syrup abuse among youths. Loose regulations, limited authority, and weak drug monitoring systems hinder prevention efforts. Despite widespread abuse, adverse effects go unreported, delaying solutions. Urgent action is needed to strengthen regulations, enhance pharmacist training, and improve reporting systems to protect youth health and improve drug monitoring (United Nations Office on Drugs and Crime, 2021).

## Recommendations

- Policy: NAFDAC should restrict OTC sales of high-risk syrups and consider prescription-only status for certain products (National Agency for Food and Drug Administration and Control, 2025).
- Practice: Provide mandatory training for pharmacists on identifying abuse and counselling youths, integrated into ongoing professional development.
- Technology: Develop user-friendly apps like MedSafety to simplify adverse effect reporting.
- Community: Launch public awareness campaigns through billboards and media, and foster collaboration with other healthcare providers and retailers.

## Questions for Further Research

- How does OTC cough syrup abuse vary across Nigeria’s regions, and what local factors influence it?
- Can user-friendly reporting apps increase pharmacists’ reporting of adverse effects?
- Does pharmacists’ financial status affect their willingness to refuse suspicious sales?
- What are the long-term health impacts of non-codeine cough syrup abuse in youths?
- How can collaboration with mental health professionals or law enforcement support pharmacists in preventing abuse?

## Data Availability

All data produced in the present work are contained in the manuscript

https://onlinelibrary.wiley.com/doi/10.1111/add.16710

